# DentalSegmentator: robust deep learning-based CBCT image segmentation

**DOI:** 10.1101/2024.03.18.24304458

**Authors:** Gauthier Dot, Akhilanand Chaurasia, Guillaume Dubois, Charles Savoldelli, Sara Haghighat, Sarina Azimian, Ali Rahbar Taramsari, Gowri Sivaramakrishnan, Julien Issa, Abhishek Dubey, Thomas Schouman, Laurent Gajny

## Abstract

**Objectives:** Segmentation of anatomical structures on dento-maxillo-facial (DMF) computed tomography (CT) or cone beam computed tomography (CBCT) scans is increasingly needed in digital dentistry. The main aim of this research was to propose and evaluate a novel open source tool called DentalSegmentator for fully automatic segmentation of five anatomic structures on DMF CT and CBCT scans: maxilla/upper skull, mandible, upper teeth, lower teeth, and the mandibular canal.

**Methods:** A retrospective sample of 470 CT and CBCT scans was used as a training/validation set. The performance and generalizability of the tool was evaluated by comparing segmentations provided by experts and automatic segmentations in two hold-out test datasets: an internal dataset of 133 CT and CBCT scans acquired before orthognathic surgery and an external dataset of 123 CBCT scans randomly sampled from routine examinations in 5 institutions.

**Results:** The mean overall results in the internal test dataset (*n* = 133) were a Dice similarity coefficient (DSC) of 92.2 ± 6.3% and a normalised surface distance (NSD) of 98.2 ± 2.2%. The mean overall results on the external test dataset (*n* = 123) were a DSC of 94.2 ± 7.4% and a NSD of 98.4 ± 3.6%.

**Conclusions:** The results obtained from this highly diverse dataset demonstrate that this tool can provide fully automatic and robust multiclass segmentation for DMF CT and CBCT scans. To encourage the clinical deployment of DentalSegmentator, the pre-trained nnU-Net model has been made publicly available along with an extension for the 3D Slicer software.

**Clinical Significance:** DentalSegmentator open source 3D Slicer extension provides a free, robust, and easy-to-use approach to obtaining patient-specific three-dimensional models from CT and CBCT scans. These models serve various purposes in a digital dentistry workflow, such as visualization, treatment planning, intervention, and follow-up.

## 1. Introduction

The clinical practice of dentistry has radically evolved in the last few years, partly because of the increasing use of digital three-dimensional (3D) data that can be gathered from dento-maxillo-facial (DMF) computed tomography (CT) scans, cone beam computed tomography (CBCT) scans, intraoral scanners, and facial scanners. This data has improved diagnosis, treatment planning, intervention, and patient follow-up in several areas of dentistry [1–4]. More specifically, patient-specific 3D models derived from CT or CBCT scans are already used for educational purposes, computer-assisted surgical planning or navigation, tooth auto-transplantation planning, and virtual treatment planning for orthodontic treatments [5–8]. It may not be long before these 3D models become a key part of precision medicine in dental practice based on finite element methods, providing an opportunity to individually assess anatomical and biomechanical characteristics and adapt treatment options accordingly [9].

To obtain a patient-specific 3D model from a CT or CBCT scan, the anatomical structures of interest must be carefully delineated on the 3D image slices, a process called segmentation. The most frequent workflows require the segmentation of jaws (maxilla and mandible), teeth (upper and lower), and the mandibular canal. When performed manually, this segmentation process takes an expert two to five hours to complete [10,11]. The current gold standard for 3D DMF image segmentation is the semi-automatic method where automatic segmentations are refined manually by an expert [12]. In recent years, several research reports have shown that deep learning-based (DL) methods could fully automate this task with results on a par with those of the experts [10,11,13–16]. Several commercially available solutions already claim to use DL methods for CBCT segmentation [17–19].

Despite these promising results, a recent systematic review of automatic tooth segmentation approaches using CBCT scans revealed that most of the studies were at high risk of bias in data selection leading to potential overestimation of the accuracy of the methods [20]. Most published studies report results from cross-validation approaches or small-sized hold-out test dataset (less than 50 CBCT scans), which is probably insufficient for evaluating the robustness and generalizability of the methods in real-world clinical settings [21].

In an effort to help the deployment and broad evaluation of rapidly evolving research, the biomedical computer imaging community has relied heavily on open research. This has led to the development of international challenges such as The Medical Segmentation Decathlon [22], the sharing of DL frameworks such as nnU-Net [23], and the sharing of pre-trained DL models for various segmentation tasks such as TotalSegmentator [24]. As far as the authors know, only two pre-trained DL models for DMF CT and CBCT segmentation are currently publicly shared. The first one is integrated in 3D Slicer software (version 5.6.1 and later - http://www.slicer.org/) [25] as an extension called Slicer Automated Dental Tools and provides segmentation of 4 anatomical structures: the mandible, the maxilla, the cranial base and the cervical vertebrae [15]. Unfortunately, this tool does not delineate the teeth from the jaws, which is a critical limitation in most digital dentistry and surgery workflows. The second one is based on the nnU-Net framework and provides segmentation of the tissues of interest, but is not supposed to be used with CBCT scan data as it has been developed and tested exclusively on CT scan data [26].

The main aim of the research presented in this paper was to propose and evaluate a novel tool for multiclass DMF CT and CBCT image segmentation called DentalSegmentator. The performance of the tool was thoroughly evaluated on two hold-out test datasets acquired from routine clinical practice in seven clinical centers.

## 2. Materials and Methods

A DL framework was trained on an internal dataset for automatic segmentation of DMF CT and CBCT scans. The results obtained from this DL-based method (the index test) were compared with those obtained by semi-automatic segmentation (the reference test) on two hold-out test datasets. The outcome set included both volume-based and surface-based metrics. The Institutional Review Board “*Comité d’Ethique pour la Recherche en Imagerie Médicale*” (CERIM) gave ethical approval for this research (IRB No. CRM-2001-051b), and its reporting conforms to recently published recommendations on artificial intelligence in dental research [21].

### 2.1. Dataset

#### 2.1.1. Patient selection

The dataset was composed of an internal dataset and an external dataset. Data from the internal dataset was selected from a retrospective sample of consecutive patients who had undergone orthognathic surgery in two French maxillofacial surgery departments. Patients referred to these public centers presented a wide variety of dentofacial deformities, came from a variety of socioeconomic backgrounds, and were ethnically diverse. Patients were considered for inclusion regardless of the dental deformity they presented, and there was no minimum age. Exclusion criteria were refusal to participate in the research and lack of industry-certified CT or CBCT scan segmentation. As a result, 603 subjects (453 CT scans, 150 CBCT scans) were included in the internal dataset.

Data from the external test dataset was randomly sampled retrospectively from routine CBCT examinations in five private centers located in India. All of the subjects were referred for a CBCT scan for various reasons such as surgical planning, orthodontic management of impacted teeth, temporomandibular joint (TMJ) disorders, or diagnosis of cysts of the jaws. Patients were considered for inclusion regardless of the condition they presented, and there was no minimum age. The only exclusion criterion was refusal to participate in the research. 123 subjects (123 CBCT scans) were included in the external test dataset.

#### 2.1.2. Data characteristics

All the scans in the internal dataset had a full-head field of view (FOV). The median in-space pixel size of the scans was 0.43*0.43mm^2^ and their median slice thickness was 0.31mm. Most CT scans (*n* = 417) were obtained using a GE Healthcare Discovery (GEHC) CT750HD scanner and all CBCT scans (*n* = 150) were obtained using a Carestream CS 9600 scanner. Scans were randomly distributed between a training/validation set (*n* = 470; 374 CT scans and 96 CBCT scans) and an internal test set (*n* = 133; 79 CT scans and 54 CBCT scans). 91% of the scans in the internal test set exhibited metal artefacts.

The FOV of the scans in the external test dataset ranged from full-head to being localized on anatomical parts (maxilla and mandible or only part of the maxilla or mandible). The median voxel size of the CBCT scans was 0.16*0.16*0.16mm^3^. The scans were acquired using five CBCT devices: Vatech Smart Plus (*n =* 25), Carestream CS 9300 (*n =* 29), Dentium Rainbow CBCT (*n =* 27), Planmeca Promax 3D (*n =* 12), and Sirona Orthophos XG 3D (*n =* 30). 42% of the CBCT scans showed metal artefacts.

Due to the anonymization process, the age of the subjects could not be retrieved. Descriptive characteristics of the dataset are shown in Table 1.

**Table 1:** Characteristics of the data in the training/validation, internal test, and external test datasets.

|  | Train/Validation | Internal Test | External Test |
| --- | --- | --- | --- |
| Number of scans | 374 CT + 96 CBCT | 79 CT + 54 CBCT | 123 CBCT |
| Median voxel size ($\text{mm}^3$) | $0.43 \times 0.43 \times 0.31$ | $0.43 \times 0.43 \times 0.31$ | $0.16 \times 0.16 \times 0.16$ |
| Number of scans by CT Device |  |  |  |
| GEHC Discovery CT 750 HD | 353 | 75 |  |
| Other CT Device | 21 | 4 |  |
| Number of scans by CBCT Device |  |  |  |
| Carestream CS 9600 CBCT | 96 | 54 |  |
| Carestream CS 9300 CBCT |  |  | 29 |
| Vatech Smart Plus CBCT |  |  | 25 |
| Dentium Rainbow CBCT |  |  | 27 |
| Planmeca Promax 3D CBCT |  |  | 12 |
| Sirona Orthophos XG 3D CBCT |  |  | 30 |
| Metal artifacts, no. (%) |  |  |  |
| Orthodontic materials | 364 (78.1) | 107 (80.5) | 3 (2.4) |
| Metal dental filling/crown | 168 (35.7) | 44 (33.1) | 49 (39.8) |
| No metal artefact | 57 (12.1) | 12 (9.0) | 71 (57.7) |
| Skeletal deformity, no. (%) |  |  |  |
| Class I | 61 (13.0) | 13 (9.8) |  |
| Class II<sup>a</sup> | 246 (52.3) | 69 (51.9) |  |
| Class III<sup>b</sup> | 163 (34.7) | 51 (38.3) |  |

<sup>a</sup>Prognathic maxilla and/or retrognathic mandible evaluated on the 3D models. <sup>b</sup>Retrognathic maxilla and/or prognathic mandible evaluated on the 3D models.

#### 2.1.3. Ground truth segmentation process (Reference Test)

The treatments carried out on patients in the internal dataset involved segmentation of the 3D scans prior to study. The ground truth segmentations were used for diagnosis, computer-aided surgical planning, and manufacture of personalized 3D-printed surgical guides and fixation implants. This was carried out by Materialise (Leuven, Belgium) according to a certified internal procedure which cannot be fully described here for reasons of confidentiality. The two-step procedure started with semi-automatic patch-based segmentations using expert priors [27] which were manually refined by an initial operator from a trained team [Step 1]. The segmentations were then verified slice-by-slice for validation by a senior operator from a trained team [Step 2] with a focus on the regions of interest (the external surface of the bones, teeth, and mandibular canals). Steps 1 and 2 were repeated until the segmentations were approved and certified for clinical use. This process resulted in five segmentation masks: maxilla/upper skull; mandible; upper teeth; lower teeth; and both mandibular canals.

The CBCT scans in the external dataset were segmented specifically for this study by using a semi-automatic three-step approach. First, the CBCT scans were segmented automatically using a publicly available deep-learning model trained on CT scans [26] [Step 1]. Second, the proposed segmentations were then corrected manually by five dentists familiar with 3D image visualization and trained for the task in 3D Slicer software (version 5.6.0) [Step 2]. Finally, the segmentations were verified slice-by-slice and corrected where necessary by a senior expert (a dentist with more than five years of experience in 3D image evaluation) in 3D Slicer software [Step 3]. This process resulted in five segmentation masks: maxilla/upper skull, mandible, upper teeth, lower teeth, and both mandibular canals.

### 2.2. Deep-Learning based segmentation (Index Test)

#### 2.2.1. Training

The nnU-Net deep learning framework (version 2.2.1) was used as an out-of-the-box tool according to instructions given by its authors [23]. Our raw training/validation internal dataset was used to automatically configure preprocessing, network architecture, and 3D full resolution U-Net training pipelines. No modifications were made in setting the nnU-Net hyperparameters and data augmentation strategy, and the target spacing of the model was 0.31*0.43*0.43mm^3^. Training time was about 24 hours on our laboratory workstation (CPU AMD Ryzen 9 3900X 12-Core; 128Gb RAM; GPU Nvidia Titan RTX 24Gb).

#### 2.2.2. Inference

Inference (prediction made by the trained model) was performed once on the internal and external test datasets following nnU-Net guidelines.

### 2.3. Evaluation

Quantitative evaluation of the model performance was carried out on the internal and external test datasets by comparing ground truth segmentations (reference test) with DL-based segmentations (index test) for each of the five segmentation masks. The recommendations of the Metrics Reloaded project [28] were followed by using both volume-based Dice similarity coefficient (DSC) and surface-based normalized surface distance (NSD). The tolerance for NSD was set at 1 mm, consistent with recent international challenges in biomedical imaging [22]. Furthermore, recent studies have shown that NSD was more strongly correlated with the amount of time needed to correct a segmentation for clinical use compared to classic metrics such as DSC [29].

### 2.4. Statistical Analysis

Continuous variables were presented as mean ± standard deviation and categorical variables were expressed as numbers and percentages. DSC and NSD results were presented as percentages (%). The results were nonparametric (Shapiro-Wilk normality test). The Wilcoxon-Mann-Whitney test was used to compare results from internal and external test datasets. The Kruskal-Wallis test was used to compare DSC and NSD results from different CT/CBCT devices; when significant, post-hoc Dunn’s test was used to compare each group. *p* values <0.05 were considered to be statistically significant. All of the data was analysed using Python (v.3.7) and R Statistical Software (v4.2.2; R Core Team 2022).

## 3. Results

### 3.1. Quantitative evaluation

Inference time was approximately 1 to 2 minutes for one 3D scan performed on the laboratory workstation described above. The mean overall results in the internal test dataset (*n* = 133) were a DSC of 92.2 ± 6.3% and an NSD of 98.2 ± 2.2% (Table 2). The mean overall results in the external test dataset (*n* = 123) were a DSC of 94.2 ± 7.4% and an NSD of 98.4 ± 3.6% (Table 3). The distribution of results is shown in Figure 1.

**Table 2:** DSC and NSD results from the internal test dataset (*n* = 133). *SD: Standard Deviation*.

| Metric, mean ± SD [%] | Maxilla / Upper Skull | Mandible | Upper teeth | Lower Teeth | Mandibular canal |
| --- | --- | --- | --- | --- | --- |
| DSC | $94.6 \pm 2.7$ | $94.5 \pm 1.6$ | $95.3 \pm 1.6$ | $95.3 \pm 1.6$ | $81.4 \pm 6.2$ |
| NSD | $97.1 \pm 2.9$ | $98.3 \pm 1.1$ | $98.9 \pm 1.0$ | $98.7 \pm 1.0$ | $98.0 \pm 3.2$ |

**Table 3:**
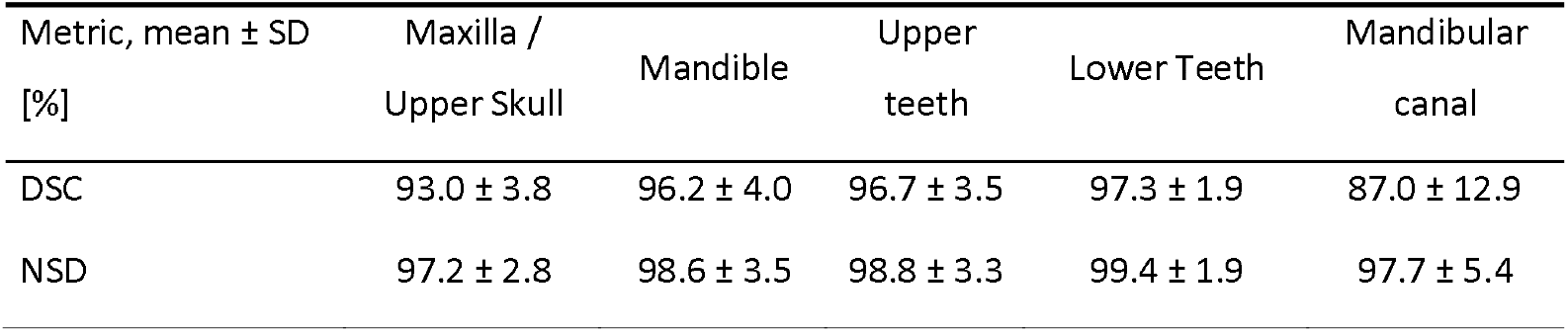
DSC and NSD results from the external test dataset (*n* = 123). *SD: Standard Deviation*.

| Metric, mean ± SD<br>[%] | Maxilla /<br>Upper Skull | Mandible | Upper<br>teeth | Lower Teeth | Mandibular<br>canal |
| --- | --- | --- | --- | --- | --- |
| DSC | 93.0 ± 3.8 | 96.2 ± 4.0 | 96.7 ± 3.5 | 97.3 ± 1.9 | 87.0 ± 12.9 |
| NSD | 97.2 ± 2.8 | 98.6 ± 3.5 | 98.8 ± 3.3 | 99.4 ± 1.9 | 97.7 ± 5.4 |

**Figure 1:**
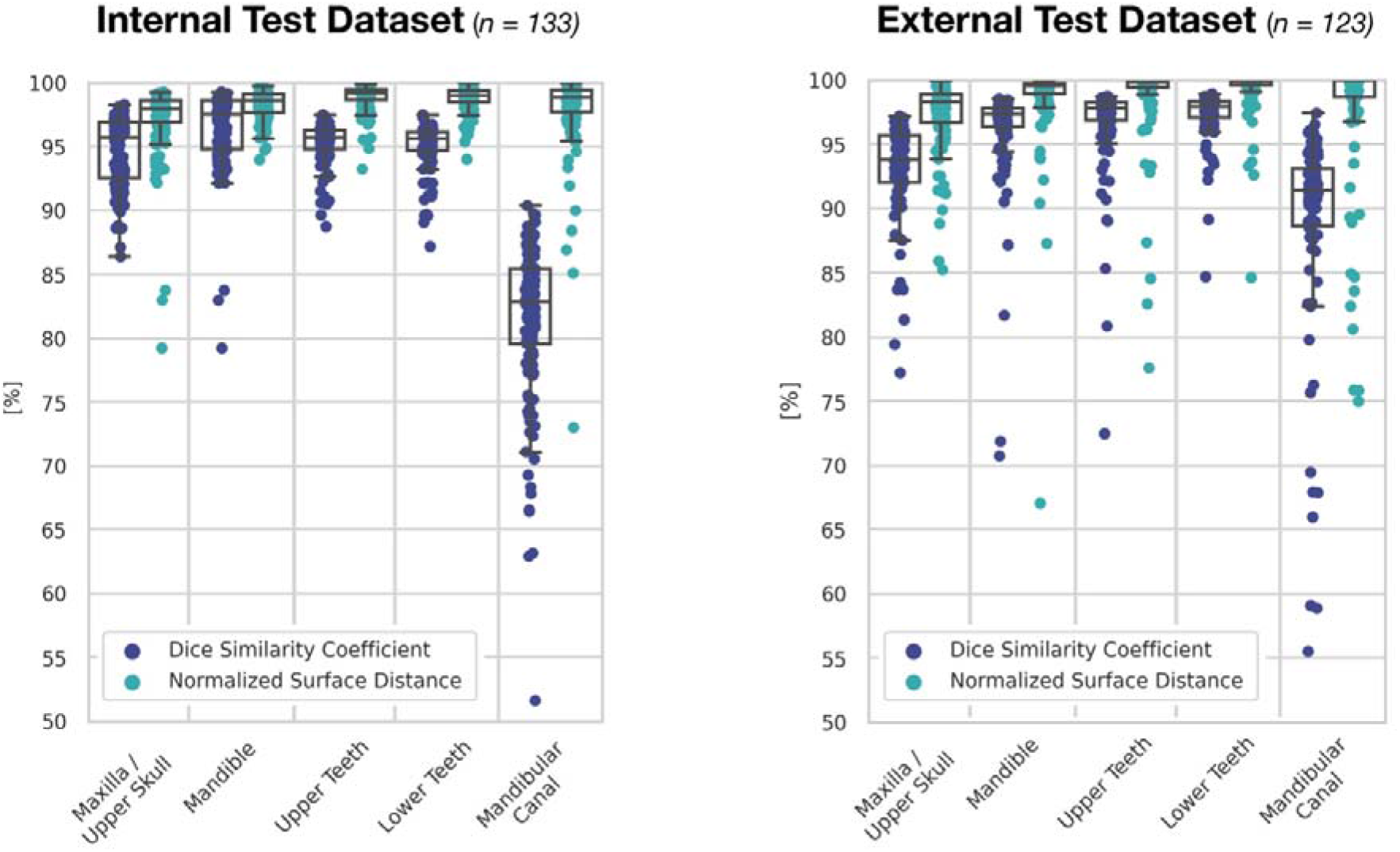
DSC and NSD results from the internal (left) and external (right) test datasets.

The statistical analysis showed similar results for both DSC and NSD metrics. Overall, the results obtained on the external and internal test dataset were statistically different. There was no statistical difference in the internal test dataset when scans obtained using the various devices were compared. In the external test dataset, the results obtained for scans acquired using Carestream 9300 and Sirona Orthophos XG 3D showed statistically significant differences.

### 3.2. Three-Dimensional Visualization

Six subjects representative of the test dataset were chosen to illustrate the segmentation results and the diversity of the CBCT data (Figure 2). When segmentation failures occurred, they were mainly under-segmentations of thin bony parts (resulting in holes in the maxilla or mandible inferior border) and missing mandibular canal parts (Figure 3).

**Figure 2:**
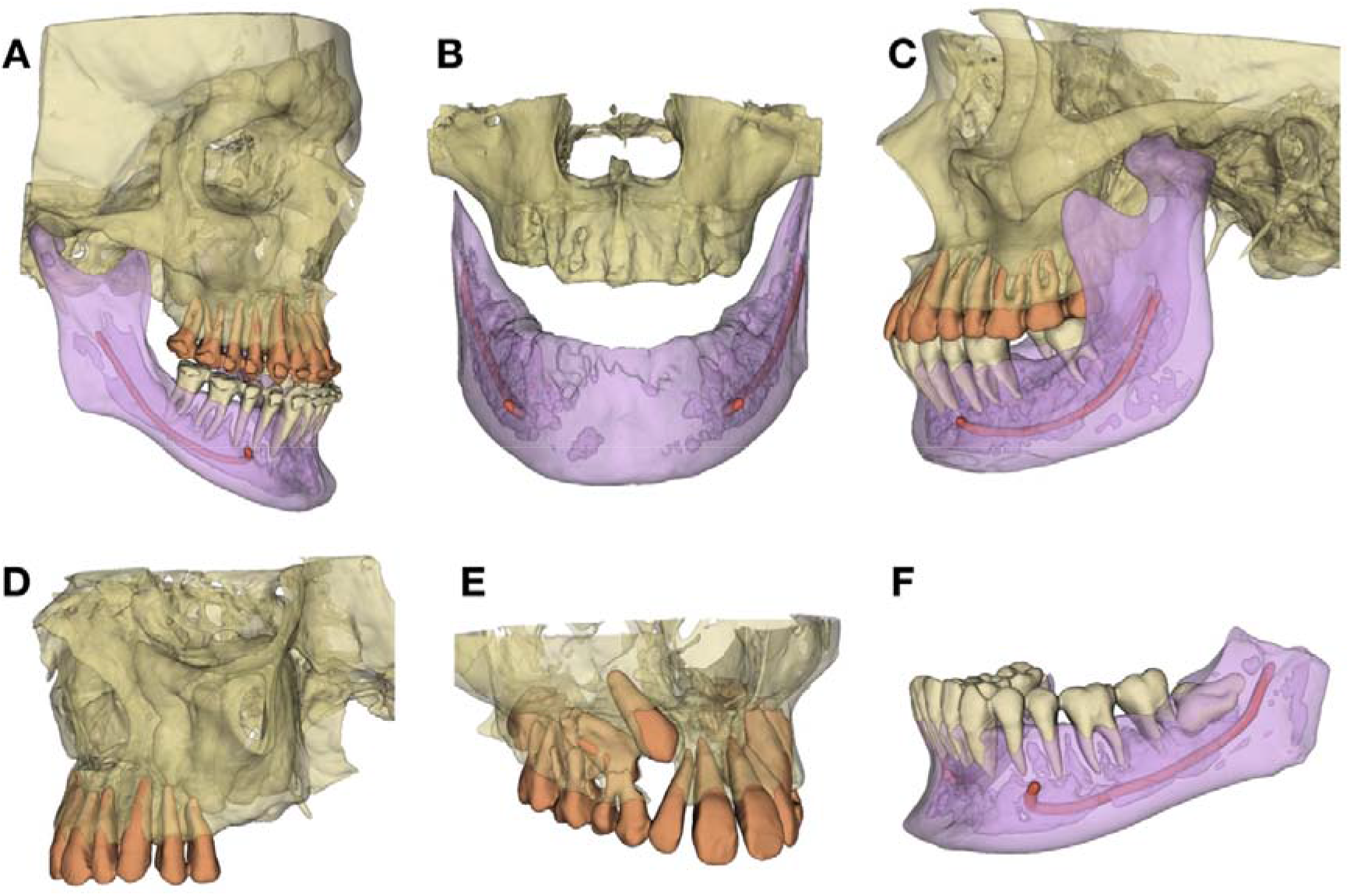
3D surface models for six subjects representative of the diversity and the challenges arising from the test CBCT dataset. (**A**) Class III maxillo-mandibular deformity, before orthognathic surgery; (**B**) Edentulous jaws; (**C**) Left condylar hyperplasia; (**D**) Upper posterior edentulous space; (**E**) Maxilla with impacted teeth; (**F**) Mandible with impacted third molar.

**Figure 3:**
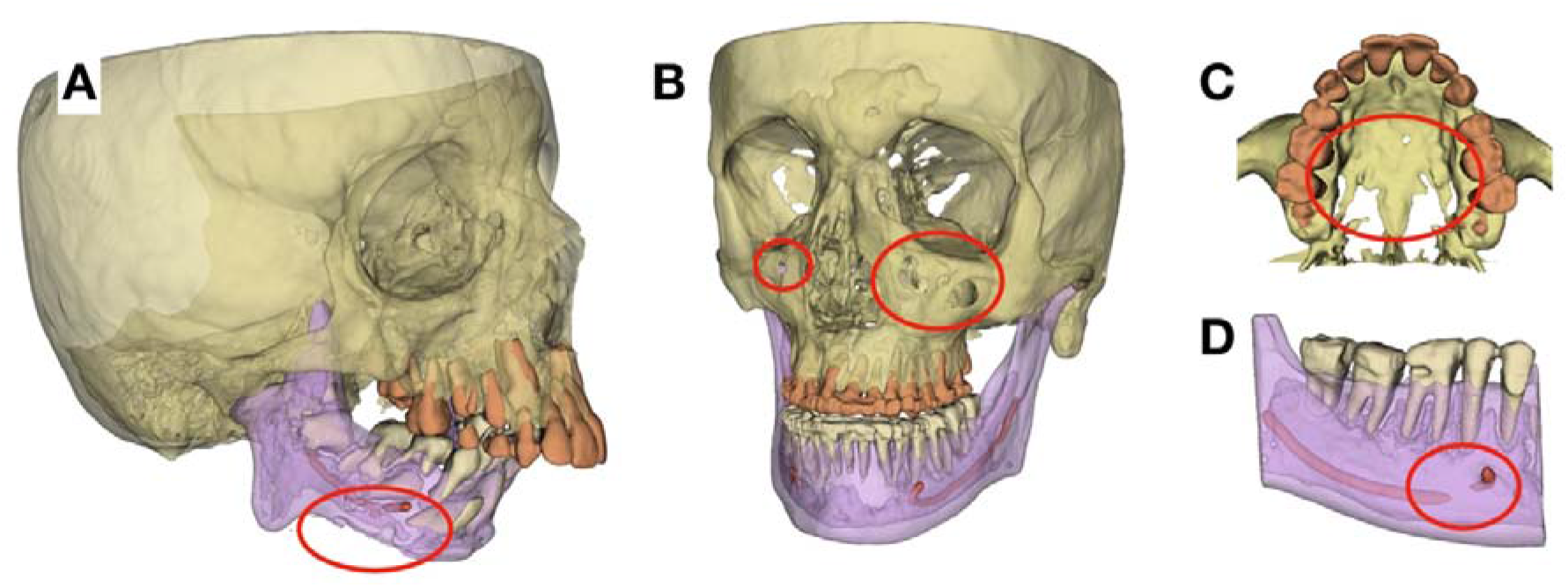
3D surface models for four subjects, exhibiting some typical failures (red circles). (**A**) Under-segmentation of the mandibular inferior border and mandibular canal; (**B**) Under-segmentation of the anterior maxillary sinus walls; (**C**) Under-segmentation of the palate; (**D**) Discontinuity of the mandibular canal.

### 3.3. DentalSegmentator model sharing and 3D Slicer extension

The pre-trained nnU-Net model is now publicly available [30]. This model can be used out-of-the-box via the nnU-Net version 2.2.1 command-line interface.

Implementation in a user-friendly interface to encourage clinicians to use the DL method is also proposed. DentalSegmentator is an open source extension for the 3D Slicer software (version 5.7.0 and later), which is a free, open source software for visualization, processing, and analysis of medical 3D images (http://www.slicer.org/) [25]. The extension, downloadable from the extension manager of 3D Slicer, offers an easy-to-use approach for DMF CT and CBCT scans automatic segmentation and 3D patient-specific model export (Figure 4). Slice-by-slice verification and manual refinement of the segmentations can be performed directly in the 3D Slicer software. More information about the extension and its code are shared on the Github platform: https://github.com/gaudot/SlicerDentalSegmentator.

**Figure 4:**
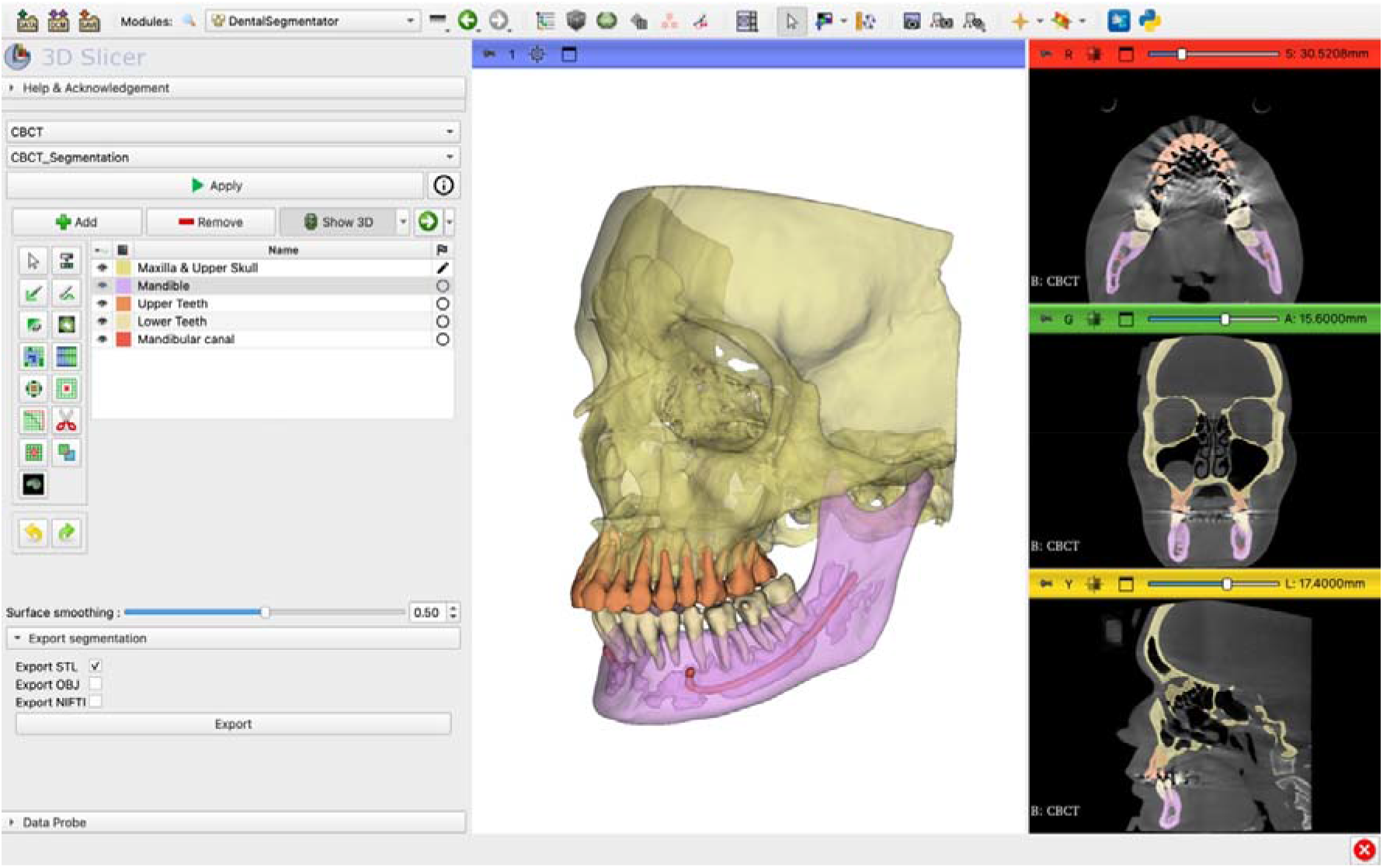
Screenshot of DentalSegmentator 3D Slicer extension.

These tools work on all computer platforms but obtaining results quickly requires a compatible graphics processing unit (GPU) with at least 4Gb of RAM. On a laptop personal computer (CPU Intel Core i7-13850HX; 32Gb RAM; GPU NVIDIA RTX 2000 8Gb), the segmentation required a mean time of 178 ± 100 seconds on 10 CT and CBCT scans randomly selected from the test dataset.

## 4. Discussion

This article introduces DentalSegmentator, a deep learning-based tool for multiclass segmentation of DMF CT and CBCT images. This tool, based on the nnU-Net framework, was evaluated on a highly diverse test dataset of 79 CT and 177 CBCT scans from seven institutions. The comprehensive evaluation was composed of both volume-based and surface-based metrics, and demonstrated that DentalSegmentator was able to provide fully automatic robust segmentation results for the five segmentation labels: maxilla/upper skull, mandible, upper teeth, lower teeth, and the mandibular canal. The pre-trained model is publicly available along with an open source 3D Slicer extension with an easy-to-use graphic interface.

Due to the lack of a publicly available DMF CT and CBCT segmentation dataset, it is difficult to compare the results obtained here directly with those previously published. A recent systematic review highlighted that the DSC results obtained in the 23 selected studies ranged from 90 ± 3% to 97.9 ± 1.5% [20]. Only a few studies in dentistry report NSC results because this metric has recently been proposed by the biomedical community [28,29]. This systematic review also pointed out that the heterogeneity in the methods employed for dataset construction and model evaluation is a frequent problem in DL studies [21]. Some of the studies excluded patients with metal artefacts or significant skeletal deformities, while most of the models were evaluated on cross-validation datasets or on hold-out test datasets of fewer than 50 CBCT scans. The main risk of these approaches is that they may yield over-optimistic results which could be difficult to reproduce in routine clinical care (i.e. poor generalizability of the model). As far as the authors know, the only study reporting results of DMF segmentation on a large-scale external test dataset (*n* = 407) had a mean DSC result of 93.8% [11], a result very close to that found in the present study. However, this model is not publicly available, which limits further evaluation and dissemination. As the dataset used in the present study was randomly selected from clinical practice, most of the test images (67.6%) showed metal artefacts. The images in our external dataset exhibited fewer metal artefacts than our internal test dataset, which might explain the statistically significant difference between the results obtained on these two datasets. Despite these statistical results, the clinical impact of the variations observed between our internal and external test results is questionable. Part of the DSC errors found in the external dataset may be caused by the resolution of the external dataset as the median voxel size of this data was half the target spacing of the model presented in this paper.

In recent years, several deep learning architectures have been proposed for the segmentation of 3D biomedical data. The study presented in this paper used the open source nnU-Net framework, which was introduced in 2018 [23]. This framework is well supported, has been shown to provide state-of-the-art results on numerous datasets, and does not necessarily require expert knowledge. New data recently published shows that in 2024 the framework was still providing highly competitive results compared to more modern architectures [30]. It is likely that slightly better results could be obtained by tailoring the deep learning architecture to the task, but that would require additional expertise.

The results demonstrated the robustness and generalizability of the model for the segmentation of routine CT and CBCT scans acquired in several cases of use such as orthognathic surgery planning, guided implant surgery, impacted teeth visualization, and digital orthodontics. Methods in health data science are evolving at a very fast pace, with growing dataset sizes and constantly improving results [32]. The results presented in this paper should improve if more training data is obtained from different CT and CBCT machines, or if post-processing steps are added (for example, to fill in some of the under-segmentations shown in Figure 3). However, automatic segmentation for DMF CT and CBCT scans is now mature enough for dental practitioners and researchers to use. This is why the pre-trained nnU-Net network and the DentalSegmentator extension for the 3D Slicer software have been publicly shared. It is hoped that this effort will help disseminate the use of 3D models in dentistry and encourage the sharing of open datasets and improved methods. It has to be said that while quantitative evaluation is necessary to assess the performance of the models, such evaluation is not always clinically relevant A clinical application such as personalized implant manufacturing will be particularly demanding in terms of segmentation precision while computer-aided diagnosis or other digital dentistry tasks may not require such precision. The more demanding the clinical situation, the more human oversight of validation and correction must be incorporated into the workflow [33].

The results presented in this paper have several limitations. The first one is their retrospective and relatively small-scale nature. A large prospective multi-center study is needed to fully evaluate the generalizability of the tool. The model was tested on 6 CBCT devices, a small number compared to the 47 CBCT devices marketed by 20 companies that were available in 2012 [34]. Moreover, it was not possible to retrieve the age of the patients in the dataset, which calls into question the applicability of the solution for subjects with primary teeth. Secondly, the construction of the reference test was a major problem due to the lack of a hard “gold standard” like dry skulls. A solid segmentation process with industry-certified segmentations (for the internal dataset) and a multi-stage approach involving experts (for the external dataset) have been provided, but bias remains possible. Finally, each tooth was not segmented and labelled separately as proposed in several other methods [11,13,16,17,19], which could be a limitation in some applications.

The ethical implications of developing and using a tool such as DentalSegmentator have not been studied at length in the literature. A checklist for the evaluation of artificial intelligence applications in dentistry from an ethical perspective has recently been proposed [35]. When evaluating the study presented in this paper from this point of view, several ethical pillars such as transparency, diversity, protection of privacy, equity, solidarity and governance were considered and addressed. More research needs to be carried out to address the pillars related to the clinical use of such a tool: wellness, respect of autonomous decision-making, accountability and responsibility, prudence, and sustainable development.

The perspectives of this study will depend on the adoption of the tool by the dental community. The main targets of this study in the short term are dental researchers and educators because 3D Slicer software is not approved for clinical use and the application distributed is intended for research use. Clinical use of this tool will require a few adaptations and further research to meet regulation such as the Artificial Intelligence Act recently adopted by the European Parliament [36]. In particular, several developments will be needed to mitigate the risk of automation bias and ensure that clinicians review and check the results before their clinical use. Thanks to the open source nature of the nnU-Net framework, the model could be easily fine-tuned with more CT and CBCT data to meet specific needs. Detection of specific pathologies like periapical lesions or bone lesions could be added to the method [37,38]. In the medium term, it is likely that other DL methods will exceed the classical 3D U-Net used in this study. For example, foundation models like the recently proposed MedSAM could allow for universal image segmentation, improving the generalizability of the current methods [39].

## 5. Conclusions

This study, based on a highly diverse dataset of more than 700 CT and CBCT scans, demonstrated the robustness of DentalSegmentator, a free open source tool for automatic segmentation of five key anatomical structures on DMF CT and CBCT scans.

## Data Availability

DentalSegmentator 3D Slicer extension and trained deep learning model are available online.

https://github.com/gaudot/SlicerDentalSegmentator

https://zenodo.org/records/10829675

